# Outbreaks of orally transmitted Chagas disease in Latin America: a comprehensive systematic review

**DOI:** 10.1101/2025.07.22.25328860

**Authors:** YM Useche, B Dinatale, OA Bottasso, AR Pérez, W Savino

## Abstract

**Background:** The rising burden of orally transmitted Chagas disease in South America underscores the need for a deeper understanding of this food-borne transmission route. Despite its relevance, key aspects such as transmission dynamics, epidemiology, and clinical outcomes remain insufficiently explored, limiting prevention and treatment strategies.

**Methods:** A systematic review (PROSPERO 2024-CRD42024542461) of reported outbreaks from 1965 to 2023 was conducted. Three reviewers independently screened and selected studies using predefined criteria. Data on epidemiology, contamination sources, clinical findings, diagnostics (cardiac, serological, molecular), vector species, and parasite lineages were extracted.

**Findings:** We compiled data from 111 outbreaks involving 1187 cases (55% males), 77 deaths, in 6 countries. Contaminated food sources were identified in 63 outbreaks (56·7%), with reliable confirmation in 8 (7·2%), *açaí* fruit being the most common source (28%). Outbreaks occurred mainly in sylvatic and tropical regions, coinciding with warm seasons and crop harvest periods. Selvatic parasite lineages predominated (98%), with *Rhodnius spp*. triatomines being implicated in 28%. The median incubation period was 22 days. Case fatality ratio was 6·5%. Acute infection presented with fever (80%), facial edema (20%), altered ECG (39·2%) and Echo (28·9%). Despite antiparasitic treatment, cardiac alterations persisted after 1 and 4 years. A ten-year follow-up showed no chronic Chagasic cardiomyopathy, though risk markers and persistent *T. cruzi*-specific IgG were reported. Data reinforces gaps mainly in long-term clinical, serological and molecular follow-up.

**Interpretation:** More integrated strategies are needed to enhance outbreak detection and management.

**Funding:** Fiocruz, CNPq and Faperj (Brazil) and FOCEM/Mercosur.

## INTRODUCTION

Chagas disease (ChD) is a potentially life-threatening illness caused by the parasite *Trypanosoma cruzi*, found from the southern U.S. to southern Argentina and Chile. In Latin America, an estimated 70 million people are at risk, with 30,000 new cases yearly, 9,000 from congenital infection, and nearly 12,000 annual deaths. The World Health Organization (WHO) prioritizes ChD among neglected tropical diseases, stressing the urgency of preventive and therapeutic advances.

*T. cruzi* has a complex life cycle involving triatomine vectors and mammalian hosts in domestic, peridomestic, and sylvatic transmission cycles. Multiple triatomine species transmit the parasite to humans and other mammals. While wild transmission mainly affects wild mammals, human cases also occur. Oral transmission is now considered to be ancestral and became a zoonosis approximately 15,000 years ago. Furthermore, *T. cruzi* is classified into 7 discrete typing units (DTUs), each linked to specific regions, vectors, clinical outcomes, and diagnostic and treatment profiles.^(1)^

The main human route of ChD infection is vector-borne transmission, followed by vertical transmission^(2–4)^, both primarily affecting rural areas where poor housing conditions favor vector infestation. Congenital cases also occur in non-endemic urban zones. However, in recent decades, numerous outbreaks of ChD seemingly caused by oral transmission have been reported in American endemic countries, showing higher morbidity and mortality than vectorial route.^(5)^ Additionally, sporadic oral ChD cases were reported in Argentina, Ecuador, and Brazil between 1936–1965.^(6–8)^ Data from DATASUS in Brazil indicate that 86.63% of acute ChD cases recorded between 2018 and 2021 were likely associated with oral transmission.^(9)^ This has led to a reconsideration of ChD as mainly vector-borne. Since 2012, WHO recognizes oral ChD as both a re-emergent and food-borne disease.^(10,11)^

ChD progresses in two phases. The acute phase lasts 2–3 months post-infection (in vector-borne and congenital cases) and is evidenced by circulating trypomastigotes in blood. Symptoms are often absent or mild and nonspecific (fever, headache, adenosplenomegaly), making diagnosis difficult. A unilateral, painless swelling of the periorbital area (Romaña sign) is pathognomonic of vectorial transmission. In contrast, oral ChD tends to cause more severe symptoms and potentially higher lethality, likely due to a larger inoculum and specific host-parasite interactions. In the chronic phase, parasites are undetectable in blood by direct methods but persist in tissues, as shown by molecular tools and reactivations in immunocompromised hosts. One to three decades after infection, ∼30% of patients may develop cardiac issues, and ∼10% can present megavisceral (megaesophagus and/or megacolon), neurological, or mixed complications.^(2,12,13)^ Benznidazole (BZN) and Nifurtimox (NFX) are the only treatments for ChD, showing effectiveness mainly during the acute phase, especially in children. Their efficacy drops in the chronic stage. A 5-year follow-up of ChD patients treated with BZN, showed a decrease in parasite detection but not a reduction in serious clinical outcomes^(14)^, despite recent findings suggesting BZN may preserve cardiac function and increase IL-17 levels in less severe chronic cases.^(15)^ Therefore, early detection and treatment of ChD appear to be critical to reduce the disease burden.

Oral ChD has become an epidemiological challenge, requiring better understanding of its development, vectors, latency, symptoms, especially those linked to lethality, and clinical follow-up. It is also crucial to study its association with geography, seasonality, and food sources to improve prevention and management strategies.^(16)^ Therefore, this study seeks to better understand the characteristics of oral ChD outbreaks through a systematic review of observational studies. In this way, we sought to outline the epidemiology and compare the characteristics of outbreaks resulting from oral ingestion of *T. cruzi* that occurred between 1965 and 2023 in several Latin American countries. Extractable outbreak data were categorized into four grades based on confidence regarding oral transmission: high, moderate, low, and very low. Finally, we also highlight the absence of crucial information in the available sources. This gap underscores the need to obtain more detailed and accurate data from patient records to establish precise protocols for outbreak reporting and management, which guide public health preventive measures for oral ChD.

This study aims to characterize oral ChD outbreaks through a systematic review of observational studies from 1965 to 2023 across Latin America. Outbreaks were categorized by confidence in oral transmission (high to very low). The lack of key data in many sources highlights the need for improved patient records to support accurate reporting and guide public health strategies.

## MATERIALS AND METHODS

### Search strategy and selection criteria

This systematic review followed the PRISMA 2020 statement (Preferred Reporting Items for Systematic Reviews and Meta-Analyses,^(17)^) and is based on a protocol (PROSPERO 2024 CRD42024542461) registered prospectively on 7 June 2024.

An electronic search for the available literature (database inception to 28 December 2023) was performed using Pubmed, Web of Science, Embase, LILACS and Scielo databases including the followed DeCS/MeSH terms: “Chagas Disease”,“*Trypanosoma cruzi* Infection”, “foodborne diseases”, “outbreak”, and other terms related to oral transmission by contaminated food like: “contaminated food”, “oral infection”, “oral transmission” and several synonyms were searched in titles and abstracts. Truncation was used to ensure variant terms were retrieved. No language limits were applied. Gray literature sources were searched by YMU and AP to identify possible relevant data about outbreaks descriptions not previously included in peer-reviewed scientific publications. These sources included PhD thesis, dissertation databases, conference abstracts, and online local newspapers reports. We also included reported cases of acute ChD from Colombian and Brazilian online government health surveillance databases. The references were exported to EndNote X7.5 and transferred to the Rayyan (https://www.rayyan.ai/) online application for the selection process.

Three authors (YMU, ARP, BD) screened titles and abstracts based on inclusion/exclusion criteria, including DeCS/MeSH terms. We excluded longitudinal, ecological, in vitro, animal, or non–Chagas-related parasitic studies. Full texts were reviewed when abstracts were inconclusive. Studies lacking diagnostic confirmation, evidence of oral infection, or extractable data were excluded. Disagreements were resolved by a fourth author (OB), following the Cochrane Handbook. Data discrepancies were discussed and checked against the original sources

### Data extraction

Data from the included studies were independently extracted by three authors (AP, YU, and BD) using an Excel form. We collected data from each outbreak, regardless of whether they involved unique or multiple outbreaks or incomplete information. Epidemiological data (date, location, country, state, city), demographic characteristics (age, sex, number of infected and deceased, latency period, and risk group), and attack rates (for exposed individuals without lab confirmation) were recorded. Diagnostic methods and clinical findings, especially cardiological alterations, were noted. We also registered data on food contamination sources, parasite and triatomine species, and the authors’ names and publication year. Authors were contacted for clarification when necessary.

### Definition of food-borne T. cruzi outbreak

An event involving at least two individuals, each with at least one laboratory-confirmed positive test for *T. cruzi* infection. Additionally, it was required the sharing of epidemiological or ChD’s clinical characteristics and/or evidence of common spatial and temporal exposure to the parasite. Outbreaks involving only one person were excluded due to difficulty in distinguishing the acute ChD resulting from the different transmission ways.

### Classification of food-borne outbreaks according to quality and quantity of extractable data

Outbreaks were categorized by the certainty level of available data:

#### High

Clinical and epidemiological evidence of acute ChD cases with temporal and spatial overlap, confirmed contaminated food source, and detection of *T. cruzi* in food and/or local triatomines. Matching *T. cruzi* DTUs in patients and vectors strongly supported oral transmission.

#### Intermediate

Clinical and/or epidemiological evidence of acute ChD cases with temporal and spatial overlap, and suspicion of contaminated food, but no *T. cruzi* detection or triatomine presence. DTU identification and exclusion of other transmission routes were lacking.

#### Limited

Clinical and/or epidemiological evidence of acute ChD cases with temporal and spatial overlap, without confirmed contaminated food, but with documented triatomines or infected animals in the area. No DTU data or exclusion of other transmission routes.

#### Scarce

Reports of temporally and spatially clustered infections with missing information on clinical features, diagnostics, food source, vector/reservoir presence, or DTU identification.

### Statistical analysis and Graph

Descriptive statistics are presented as frequencies for categorical variables, means and standard deviations for normally distributed data, or median with range or quartiles for other continuous variables. Normality was assessed using the Shapiro-Wilk test. Categorical variables were compared using the chi-square test or Fisher’s exact test when appropriate, while continuous variables were analyzed using Student’s t-test or the Mann-Whitney U test, depending on data distribution. The geographical distribution map was created using Scimago Graphica (https://www.graphica.app/), incorporating geolocation data obtained from bibliographic sources or Google Maps.

## RESULTS

The database search yielded 797 reports (including observational studies as outbreak reports, outbreak series or reviews with details of outbreak series, as well as studies of molecular typing of the strains involved in the outbreaks) and 42 grey literature articles (including theses, conference abstracts, governmental health agency reports, and online journals, while press reports were included to analyze seasonality). After automatic removal for duplication (474) and for ineligible reports (209) including longitudinal, experimental, and animal studies, 121 reports were searched for retrieval, but only 114 peer-reviewed and 36 grey literature articles were retrieved. After undergoing a title and abstract screening, searching for studies without oral infection or isolated case reports, 107 peer-reviewed reports and 36 grey literature articles were selected for full-text review. Publications were then excluded due to the absence of extractable data or information on positive diagnostic tests. As seen in **Figure 1**, finally, a total of 110 reports corresponding to 111 outbreaks were incorporated in this systematic review, which included 1187 cases of orally-transmitted acute Chagas disease that occured from 1965 until December 2023 (**Table 1 and Figure 2**)

**Figure 1.**
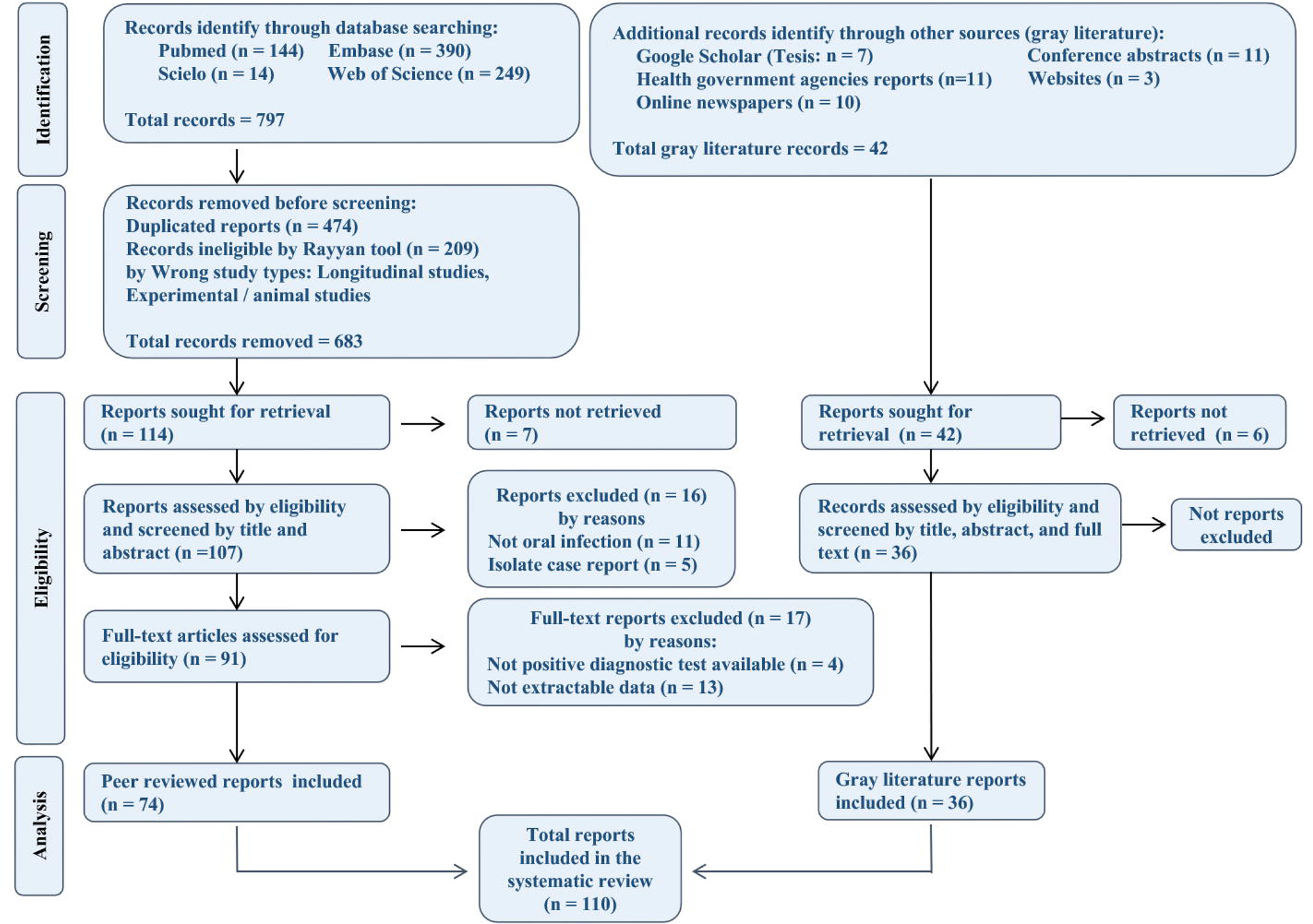
Preferred reporting items for the systematic review flowchart of the study selection process.

**Figure 2.**
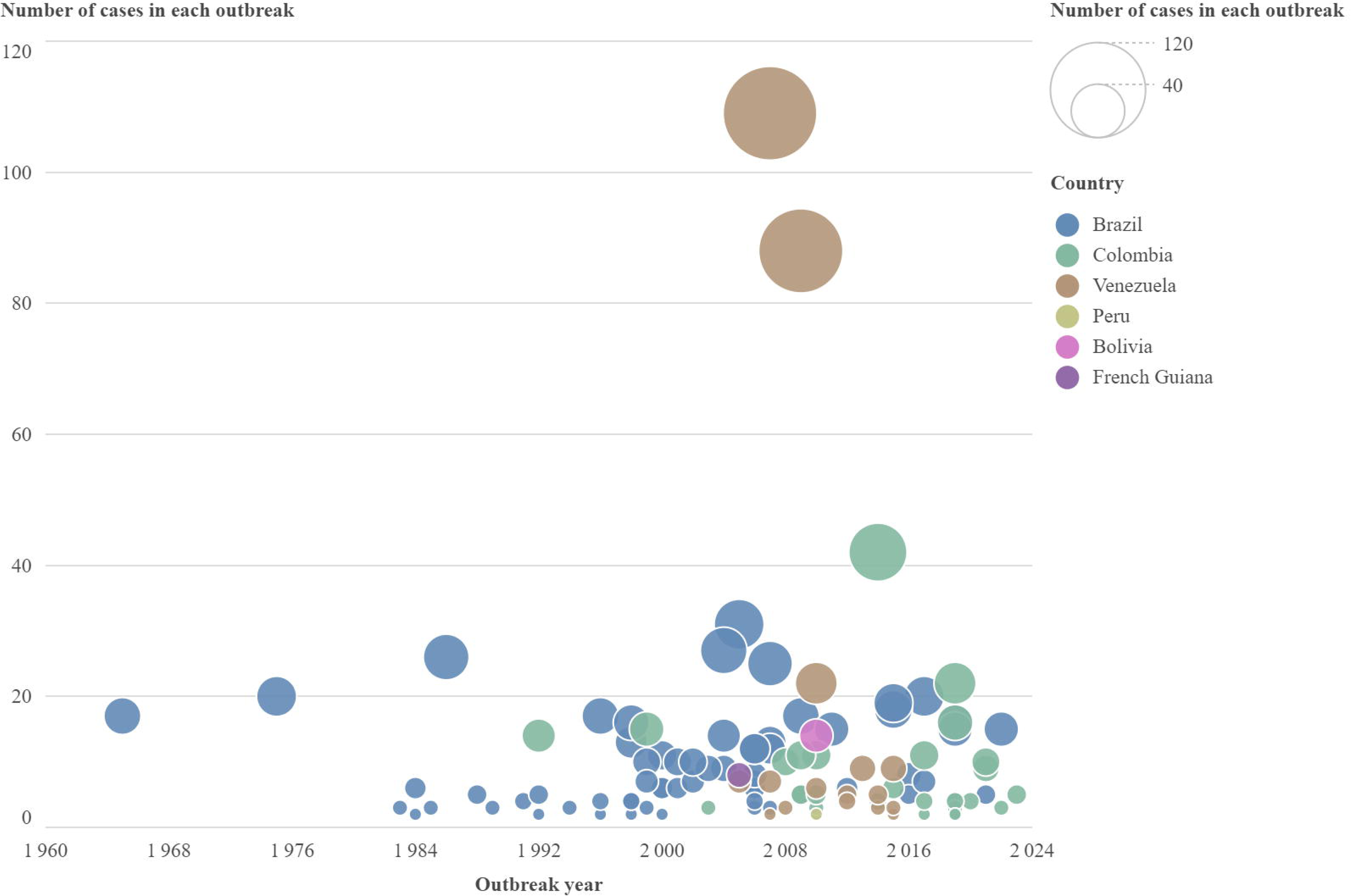
Annual number of orally transmitted Chagas disease outbreaks and associated cases per outbreak in Latin American countries, 1965–2023. Note: Although the first outbreak was officially reported in 1998, it retrospectively occurred in 1965. The relative size of each outbreak, in terms of reported cases, is represented by the diameter of the circles.

**Table 1.** General features of oral ChD outbreaks occurred in the period of 1965 to 2023.

| Country | Outbreaks (n) | Cases (n) | Sex distribution F/M/(N.a.) | Age of oral ChD cases (mean -years-± SD) | Total Deaths | Death distribution F/M/(N.a.) | Age of deaths (mean, - years- ±SD) | CFR (%)<sup>(*)</sup> | Attack rate (%)-[CI] |
| --- | --- | --- | --- | --- | --- | --- | --- | --- | --- |
| **Bolivia** | 1 | 14 | N.a. | N.a. | 0 | 0/0 | - | 0 | N.a. |
| **Brazil** | 62 | 628 | 142/156 (330) | 26·1 ± 10·1 | 29 | 8/14 (7) | 23·7 ± 23·5 | 4·79 | 43·5<br>[95% CI 55·2-31·7] |
| **Colombia** | 30 | 249 | 46/138(65) | 27·2 ± 10·9 | 23 | 1/4 (21) | 19·2 ± 18·9 | 8·7 | 58·7<br>[95% CI 75·6-41·7] |
| **French Guiana** | 1 | 8 | N.a. | 36·1 ± 0 | N.a. | N.a. | N.a. | - | 100 |
| **Peru** | 1 | 2 | 0/2 | 3·0 ± 0 | 0 | 0/0 | - | 0 | 50 |
| **Venezuela** | 16 | 286 | 124/111 (51) | 23·0 ± 11·9 | 25 | 6/8 (15) | 20·5 ± 23·5 | 8·59 | 83·5<br>[95% CI 109·4-57·6] |
| **Total** | 111 | 1187 | 312/389 (486) | 25·5 ± 11·0 | 77 | 15/26 (43) |  | 6·7 | 55·3%<br>[95% CI 64·4-46·1] |

**ChD:** Chagas disease; **F:** Female; **M:** Male; **n:** numbers, **N.a.:** not available. <sup>(\*)</sup> **CFR: Case fatality rate** was calculated as the total reported deaths by oral CD infection/total number of diagnosed cases of oral ChD infection. **SD:** Standard Deviation

As seen in **Figure 2**, the number of reported food-borne ChD outbreaks that fulfill the inclusion criteria was 14 in the period between 1965 and 1995. However, from 1996 to 2023, this number increased to 97, showing almost a 7-fold increase in reported outbreaks over the same period of time (27 years). The highest number of reported outbreaks in order was in Brazil, Colombia and Venezuela, the country that exhibited the 2 largest outbreaks in terms of absolute number of cases (88 and 109 cases).

The geographical distribution of outbreaks and their seasonality provide key insights into how environmental factors may influence oral transmission dynamics. As depicted in the georeferencing map (**Figure 3**), food-borne ChD outbreaks are predominantly concentrated in sylvatic or tropical regions (mainly within the Amazon basin) or in subtropical areas associated with agricultural activities. Notably, outbreaks often coincide with the harvesting periods of tropical fruits and crops, such as sugarcane in Santa Catarina, Brazil, suggesting a potential link between agricultural practices and the risk of oral transmission.

**Figure 3:**
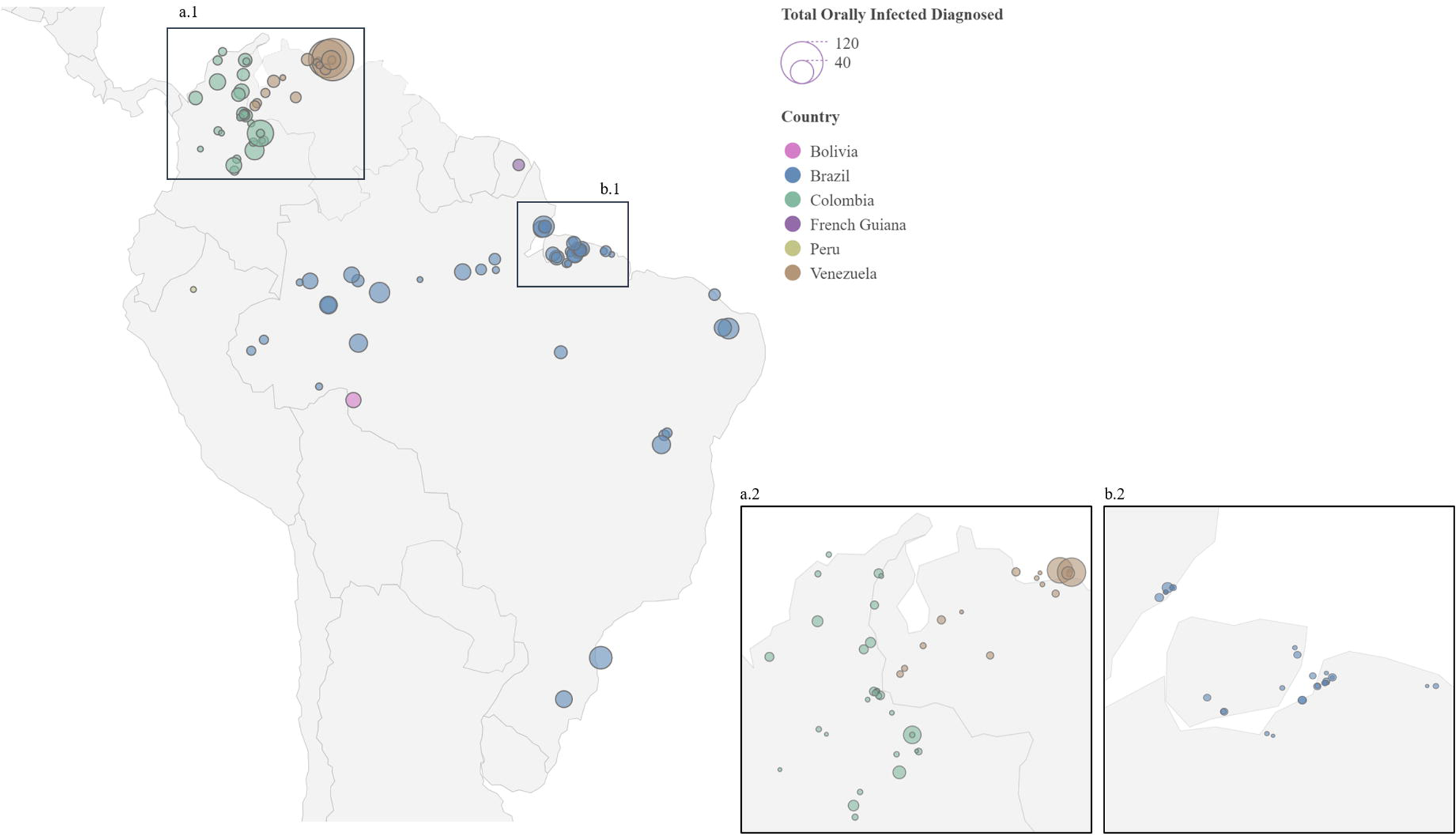
Geolocation of food-borne Chagas disease outbreaks that occurred between 1965 and 2023. The areas highlighted in **a1** and **b1** are shown in greater detail in **a2** and **b2**, respectively. Associated cases per outbreak are also shown.

Analyzing the seasonality of outbreaks can provide valuable insights into the dynamics of food-borne transmission of *T. cruzi*. Seasonality data were available for 82 out of the 111 outbreaks (73·9%). As shown in **Figure 4**, outbreaks usually occur during the warmest and wettest months of the year reported for these areas. The period from March to May experienced the highest concentration of outbreaks, with 30 outbreaks and 383 cumulative cases, followed by the October to December period with 23 outbreaks and 358 cumulative cases. In contrast, only 3 outbreaks were reported in July, yielding 29 cumulative cases.

**Figure 4:**
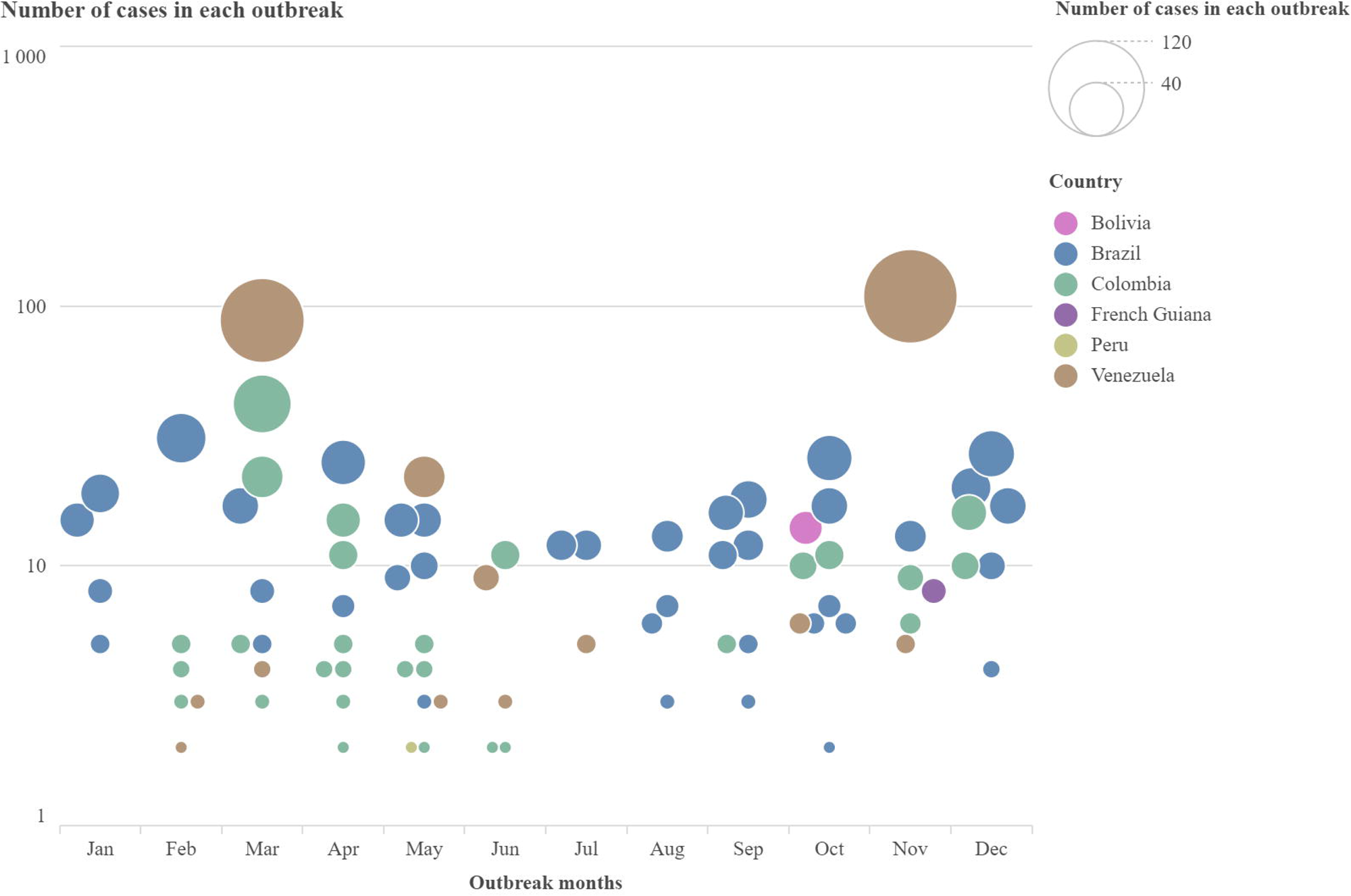
Seasonality of orally transmitted Chagas Disease outbreaks occurred between 1965-2023. Associated cases per outbreak are also shown.

Only a few studies have provided complete information on the outbreak’s epidemiological curve. We extracted or reconstructed 19 outbreaks (17·1%) in which the onset of symptoms following oral ingestion of *T. cruzi*-contaminated food or beverages varied between 2- and 25-days post-exposure, with the median incubation period being 16 days.

In 62 outbreaks (55·4%), reports identified the exposed populations, involving relatives in about half of these (55·5%), coworkers (11·6%), groups that consumed commercial food (7·1%), neighbors or non-relatives (6·3%), and members of closed communities such as schools (n=4) or elderly care facilities (n=1; 4·5%). In 15 outbreaks, the affected groups included more than one category (i.e., relatives and non-relatives).

Based on 60 reports with reliable sex and/or age distribution data, the mean proportion of infected females was 44.5% (n=312) and males was 55·5% (n=389), while the mean age of infected individuals was 25·5 (ranging from 1 to 80 years old). In most of the outbreaks, neither the sex nor the age of the involved people was reported (**Table 1**). The total reported deaths during the 1965-2023 period were 77, whereas the case fatality rate (reported as number of cases/total reported deaths) was 6·52%. The global attack rate was 56%. A sub-analysis based on the country distribution of oral ChD outbreaks showed significant differences in attack rates among countries with the highest number of cases, varying from 43·5% in Brazil to 83·5% in Venezuela (where the attack rates for the two highest reported outbreaks are unavailable) (**Table 1**). Nine pregnant women orally-infected were found, 5 of them experiencing abortions, representing a 55·5% fetal loss.

Laboratory assays are crucial for confirming *T. cruzi* as the causal agent of outbreaks. However, out of the reports included in this systematic review, only 89 (80·2%) provided thorough information on the laboratory methods of diagnosis used in each outbreak. Among the total number of laboratory diagnosed patients (n=1153/1187) by one or more major methods (parasitological, serological or molecular), detailed methods and positive reports were only reported in 88·4% of them (n=1019/1153).

The quality of diagnostic tests for ChD has significantly evolved from 1965 to the present.^(18,19)^ Reports from outbreaks occurring from 1965 until 2023 showed that, globally, direct microscopic observation (Strout/microStrout, wet smear) yielded similar number of positives (41·8%, n=480) than serological methods, including ELISA, IFI and HAI, which showed the highest diagnostic yield, being reactive in 68·6 % of infected patients (n=699). Additional parasitological methods such as culture (17·2%; n=201) and xenodiagnosis (17·2%, n=197) were chosen as diagnostic methods in the 1960-1980 decades. Other techniques were also used to diagnose 143 patients (12·5%). Among them, there were inoculation of laboratory animals (5 outbreaks), complement mediated lysis (2 outbreaks), direct examination of pericardial fluid or cerebrospinal fluid (2 outbreaks), Tesa blots (2 outbreaks) and immunoprecipitation in one case. Biopsy and necropsy were also utilized to diagnose acute oral ChD (in 4 outbreaks). More recently, the implementation of molecular methods (PCR/qPCR), allowed or reinforced the diagnosis in 248 subjects (21·6%). In the outbreak reports included here, there was no mention of the use of more recently developed diagnostic tests, such as chemiluminescent magnetic immunoassays, loop-mediated isothermal amplification (LAMP) or rapid diagnostic tests.^(20)^ **Table 2** depicts the frequency of orally infected subjects diagnosed with parasitological, serological, molecular and other methods used along the analyzed period (Note that a patient can be diagnosed using more than one major method).

**Table 2:** Frequency of subjects diagnosed with different laboratory techniques during oral ChD outbreaks.

| Diagnostic Techniques (DT) | Number of patients with positive tests | Overall Positivity Rate (%)* | Test-Specific Positivity Rate (%)** |
| --- | --- | --- | --- |
| **Parasitologic Tests** | **569** | **55.8** |  |
| -Direct | 480 | 47.1 | 59.6 |
| -Xenodiagnosis | 197 | 19.3 | 69.4 |
| -Culture | 252 | 24.7 | 54.0 |
| **Serologic Tests** | **699** | **68.6** |  |
| -IHA | 422 | 41.4 | 75.6 |
| -ELISA<br>- IgG<br>- IgM<br>- Non informed isotype | 379 | 37.2<br>28.6<br>26.0<br>8.4 | 84.4 |
| -IFI<br>- IgG<br>- IgM<br>- Non informed isotype | 385 | 37.7<br>27.1<br>25.5<br>7.4 | 82.7 |
| **Molecular Tests (PCR/qPCR)** | **248** | **24.3** | **52.3** |
| **Other** | **143** | **14.0** | **37.7** |

\*Overall Positivity Rate (%) = (number of positive tests/ 1019 patients x 100)
\*\*Test-Specific Positivity Rate (%) = (number of positive tests/laboratory diagnosed patients in each outbreak) \*100

When we consider the total number of infected individuals included in this systematic review (n=1187), clinical manifestations were mentioned in 914 patients, but detailed individualized reports were available for only 64·9% of them (n=771, depicted in **Figure 5a**). Based on these reports, it was evident that most acutely infected patients presented prolonged fever (79·9%, n=616), whereas other manifested systemic symptoms, including myalgia (35·3%), headache (35·0%) and facial edema (22·7%). Other less frequent symptoms were also described, like lower limb edema, fatigue, anasarca, abdominal pain, dyspnea, arthralgia, hepatomegaly, skin signs (as cutaneous erythematous rash or exanthems), vomiting, bipalpebral and bilateral edema, splenomegaly, lymphadenopathy, epigastralgia, chest pain, anorexia, cough, malaise, upper limb edema and meningoencephalitis (2 cases). Diarrhea, hemorrhagic manifestations (epistaxis, hematemesis, melena), jaundice, and palpitations were also occasionally mentioned. Electrical and echocardiographic studies were detailed reported for 474 patients (61·5%), with alterations detected in 186 of them (39·2%). Echocardiographic abnormalities were present in 137 (28·9%) infected subjects, with 54 (11·4%) of them developing both electrical and echocardiographic abnormalities. Of the total number of deaths, cardiac disturbances were listed in only 26 cases, one case of acute myocardial infarction and 9 necropsies confirming myocarditis. Main cardiac disturbances are summarized in **Figure 5b**. Depolarization disorders, tachycardia, pericardial effusion, heart failure, branch blocks, and pericarditis were the most common findings during the acute phase of ChD. Evidence associated with a poor prognosis was also recorded, such as complete right bundle branch block, extrasystoles, sinus tachycardia, and atrial and ventricular fibrillation. Segmental wall motion abnormalities (hypokinesis, akinesis, or dyskinesia) were also frequent.

**Figure 5.**
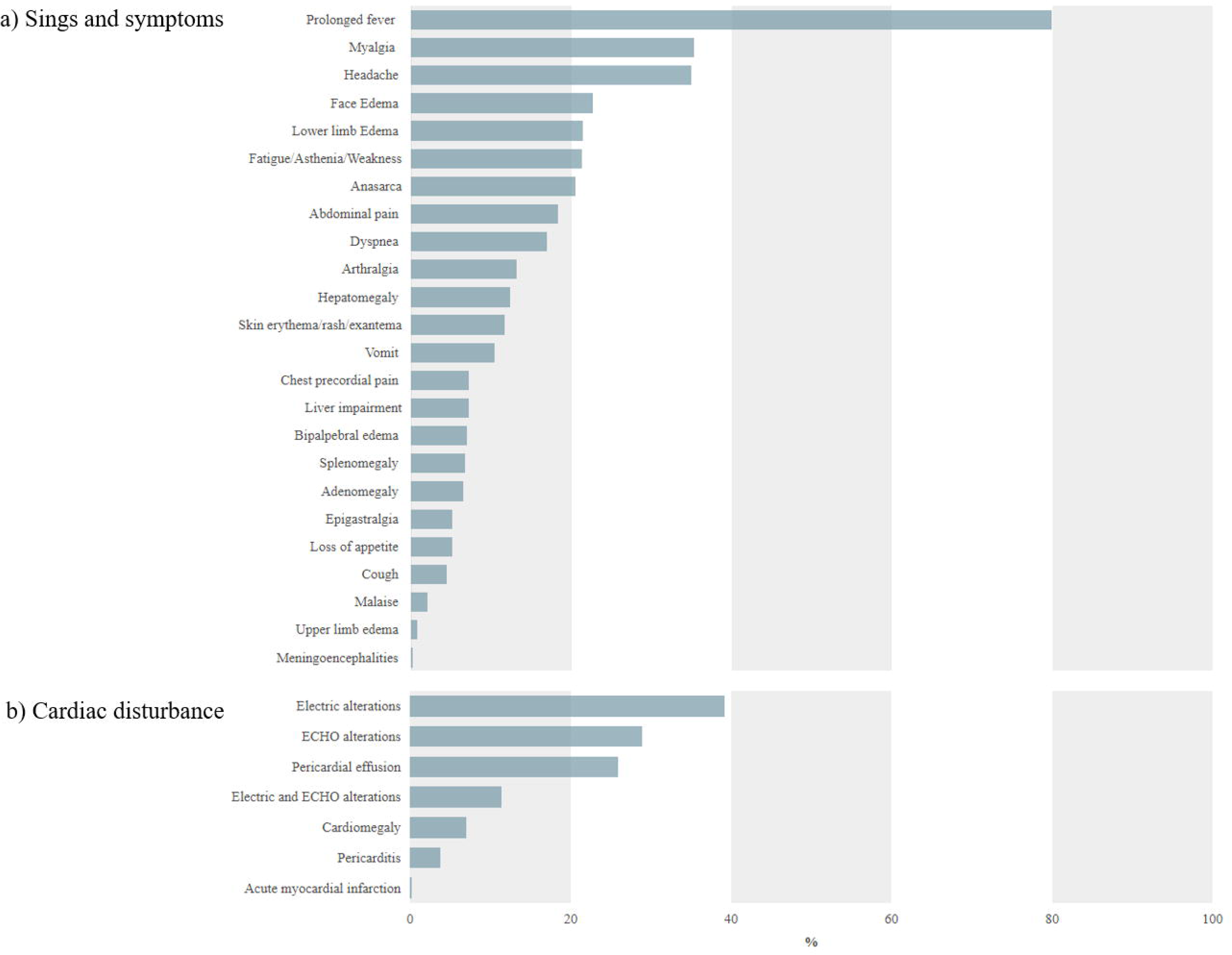
(**a**) Frequency of signs and symptoms in 771 patients with food-borne ChD. (**b**) Major cardiac disturbances reported in 493 cardiologically evaluated patients.

Despite that, cardiological assessments performed by ECG, Holter, or echocardiography, were detailed reported in only 474 cases (61·5%) of orally infected patients, corresponding to 54 outbreaks (**Table 3**). Among them, pericardial effusion was detected in 123 patients (25·9%), and cardiomegaly in 33 patients (6·9%).

**Table 3:** Detailed electro and echocardiographic abnormalities detected in *T. cruzi*-orally infected patients with cardiological evaluation.

|  | Reported Cases<br>(n)(*) | Reported<br>Alterations<br>(n)(**) | % |
| --- | --- | --- | --- |
| **Total number of evaluated patients** | **474** |  | **100.0** |
| **Repolarization disorders** | **132** |  | **27.8** |
| -Without description |  | 57 | 12.0 |
| -Other Repolarization Disorders |  | 37 | 7.8 |
| -Disturbances in Ventricular Repolarization |  | 27 | 5.7 |
| -Disturbances in Auricular Repolarization |  | 11 | 2.3 |
| **Pericardial effusion** | **123** |  | **25.9** |
| **Tachycardia** | **83** |  | **17.5** |
| -Sinus tachycardia |  | 34 | 7.2 |
| -Without description |  | 32 | 6.8 |
| -Atrial tachycardia |  | 14 | 3.0 |
| -Ventricular tachycardia |  | 3 | 0.6 |
| **Branch Blocks** | **41** |  | **8.6** |
| -AV block |  | 21 | 4.4 |
| -Complete right bundle branch block |  | 15 | 3.1 |
| -Anterosuperior hemiblock |  | 6 | 1.3 |
| -Right bundle branch hemiblock |  | 3 | 0.6 |
| -Complete left bundle branch block |  | 7 | 1.5 |
| **Heart failure** | **36** |  | **7.6** |
| **Pericarditis** | **18** |  | **4.0** |
| **Mildly reduced or reduced Ejection Fraction (<49%)** |  | 16 | 3.4 |
| **Mitral Failure and Regurgitation** |  | 12 | 2.5 |
| **Atrial Fibrillation (AF)** |  | 12 | 2.5 |
| **Ventricular hypertrophy** |  | 11 | 2.3 |
| **QRS low voltage (LV)** |  | 11 | 2.3 |
| **Extrasystoles** |  | 9 | 1.9 |
| **Aneurysmal dilations** |  | 2 | 0.4 |
| **Sinus Bradycardia (SB)** |  | 5 | 1.1 |
| **Syncope** |  | 1 | 0.2 |

The data are presented as follows: (\*) the total number of patients with abnormalities, and (\*\*) the total number of abnormalities detected among patients when the exact number of affected individuals is not specified. In the latter case, it is not possible to determine whether multiple abnormalities occurred in the same individual, preventing direct correlation with patient numbers.

Cardiological follow-up reports were available for only 14 outbreaks (12·2%, 14/111), with all included patients subjected to antiparasitic treatment. There were no reports of megacolon or megaesophagus cases (nor was it explicitly stated that its presence had been evaluated). Regrettably, cardiological assessments were not consistently included in the reports for patients comprised within each outbreak. In sum, 160 individuals underwent different schedules of cardiological follow-up, representing a small proportion of the overall infected population (13·5%, 160/1187), even when including those with cardiological evaluations conducted during the acute phase. The timing of follow-up assessments varied considerably, ranging from assessments conducted in the months following antiparasitic treatment completion to as long as 10 years post-infection

As seen in **Table 4**, the monitoring of cardiac abnormalities in the early post-treatment period (follow-up between treatment completion and 6 months) revealed that 40·4% (40/99) exhibited persistent electrocardiographic abnormalities, while none of the evaluated patients (n=29) showed echocardiographic alterations. The proportion of electro and echocardiographic abnormalities was significantly lower than that seen during the acute phase (p=0·0001, Fisher’s exact test in both cases). In the follow-up period between 6 months and 1 year, electrocardiographic abnormalities remained noticeable, with incomplete right bundle branch block (18/114), sinus tachycardia (ST; 16/114), and T-wave inversion (15/114) being the commonest findings, followed by ventricular repolarization dispersion (6/114), extrasystoles (5/114), and atrial tachycardia (AT; 2/114). In the same period, echocardiographic assessments, available for a smaller subset of patients, indicated mitral regurgitation (3/23), abnormal relaxation (1/23), pericardial effusion (1/23), and diffuse hypokinesis (1/23).

**Table 4:**
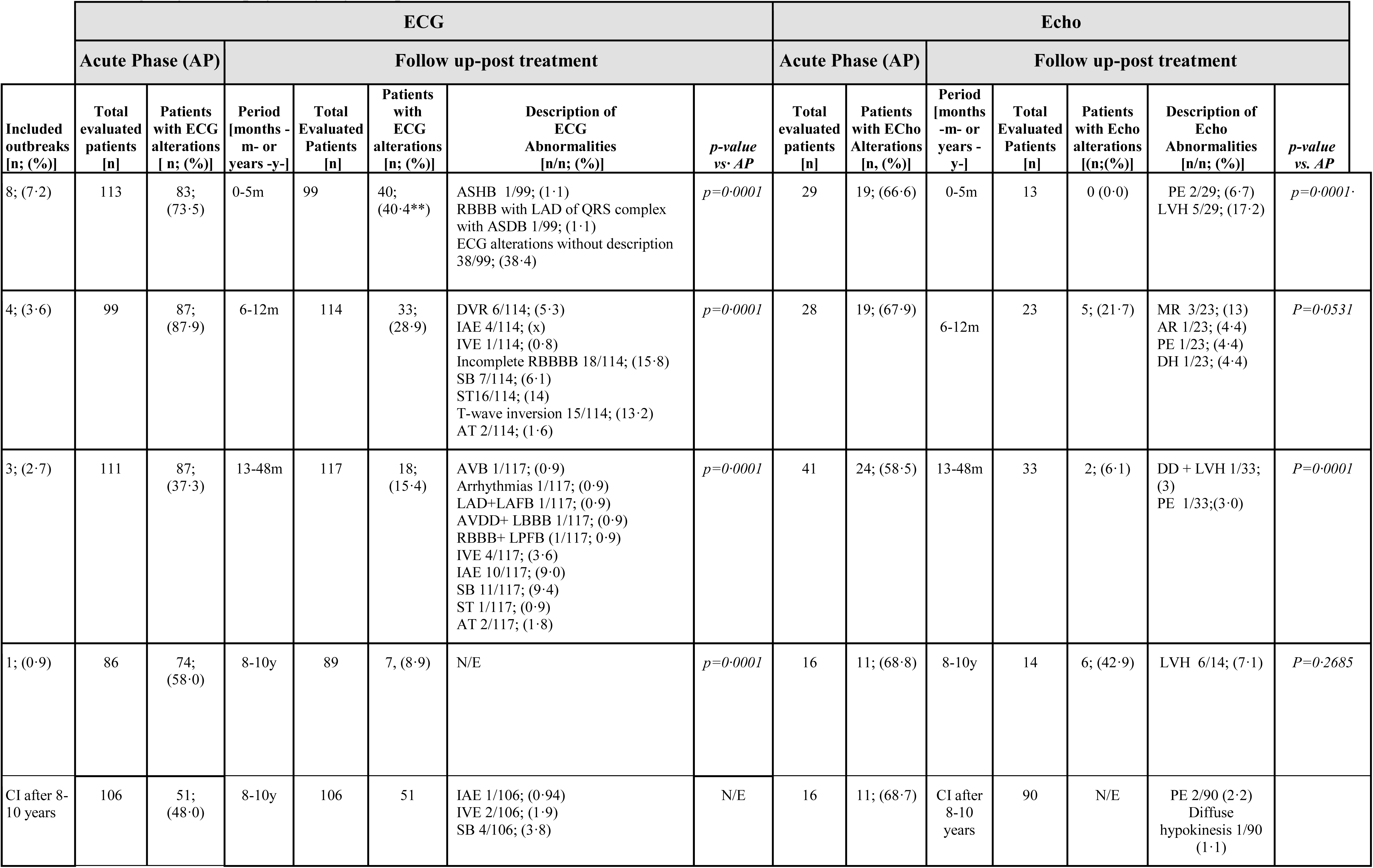
Cardiological follow-up of orally-infected patients.

|  | ECG |  |  |  |  |  |  | Echo |  |  |  |  |  |  |
| --- | --- | --- | --- | --- | --- | --- | --- | --- | --- | --- | --- | --- | --- | --- |
|  | Acute Phase (AP) |  | Follow up-post treatment |  |  |  |  | Acute Phase (AP) |  | Follow up-post treatment |  |  |  |  |
|  | Total evaluated patients<br>[n] | Patients with ECG alterations<br>[ n; (%)] | Period [months - m- or years -y-] | Total Evaluated Patients<br>[n] | Patients with ECG alterations<br>[n; (%)] | Description of ECG Abnormalities<br>[n/n; (%)] | *p-value vs. AP* | Total evaluated patients<br>[n] | Patients with ECHO Alterations<br>[n, (%)] | Period [months -m- or years -y-] | Total Evaluated Patients<br>[n] | Patients with Echo alterations<br>[(n;(%)] | Description of Echo Abnormalities<br>[n/n; (%)] | *p-value vs. AP* |
| 8; (7.2) | 113 | 83; (73.5) | 0-5m | 99 | 40; (40.4**) | ASHB 1/99; (1.1)<br>RBBB with LAD of QRS complex with ASDB 1/99; (1.1)<br>ECG alterations without description 38/99; (38.4) | *p=0.0001* | 29 | 19; (66.6) | 0-5m | 13 | 0 (0.0) | PE 2/29; (6.7)<br>LVH 5/29; (17.2) | *p=0.0001* |
| 4; (3.6) | 99 | 87; (87.9) | 6-12m | 114 | 33; (28.9) | DVR 6/114; (5.3)<br>IAE 4/114; (x)<br>IVE 1/114; (0.8)<br>Incomplete RBBBB 18/114; (15.8)<br>SB 7/114; (6.1)<br>ST16/114; (14)<br>T-wave inversion 15/114; (13.2)<br>AT 2/114; (1.6) | *p=0.0001* | 28 | 19; (67.9) | 6-12m | 23 | 5; (21.7) | MR 3/23; (13)<br>AR 1/23; (4.4)<br>PE 1/23; (4.4)<br>DH 1/23; (4.4) | *P=0.0531* |
| 3; (2.7) | 111 | 87; (37.3) | 13-48m | 117 | 18; (15.4) | AVB 1/117; (0.9)<br>Arrhythmias 1/117; (0.9)<br>LAD+LAFB 1/117; (0.9)<br>AVDD+ LBBB 1/117; (0.9)<br>RBBB+ LPFB (1/117; 0.9)<br>IVE 4/117; (3.6)<br>IAE 10/117; (9.0)<br>SB 11/117; (9.4)<br>ST 1/117; (0.9)<br>AT 2/117; (1.8) | *p=0.0001* | 41 | 24; (58.5) | 13-48m | 33 | 2; (6.1) | DD + LVH 1/33; (3)<br>PE 1/33;(3.0) | *P=0.0001* |
| 1; (0.9) | 86 | 74; (58.0) | 8-10y | 89 | 7, (8.9) | N/E | *p=0.0001* | 16 | 11; (68.8) | 8-10y | 14 | 6; (42.9) | LVH 6/14; (7.1) | *P=0.2685* |
| CI after 8-10 years | 106 | 51; (48.0) | 8-10y | 106 | 51 | IAE 1/106; (0.94)<br>IVE 2/106; (1.9)<br>SB 4/106; (3.8) | N/E | 16 | 11; (68.7) | CI after 8-10 years | 90 | N/E | PE 2/90 (2.2)<br>Diffuse hypokinesis 1/90 (1.1) |  |

Studies conducted between the first and fourth year of follow-up revealed a series of conduction and rhythm disturbances in 18/117 patients. Among these, predominated sinus bradycardia (SB; 9·4%), isolated atrial extrasystoles (IAE; 9%), isolated ventricular extrasystoles (IVE; 3·6%) and AT (1·6%), with other alterations being recorded in a small proportion (<1% per alteration). Moreover, diastolic dysfunction, left ventricular hypertrophy and pericardial effusion were detected by Echo in 1/33 patients (3%).

A long-term follow-up study, that is 8–10 years post-infection corresponded to the Chacao outbreak wherein 10·1% (9/89) patients persisted with abnormalities in either electro or echocardiographic surrogates(21). Furthermore, radiographic imaging indicated that 10·8% of evaluated patients presented a cardiothoracic ratio >0·5 and left ventricular atrophy (1/14, 7·1%). In this outbreak, many patients received a second treatment with BZL, due to parasite persistence. However, none of these alterations could be regarded as chronic cardiomyopathy, despite that certain findings, such as SB in 4 patients, may represent early manifestations of the disease.

As seen in **Table 5**, long-term follow-up evaluations of oral Chagas disease were scarce in terms of seronegative conversion or parasite persistence: 10·8% (12/111) and 9% (10/111), respectively. The 60-day regimen of BZL was the most recommended treatment in these outbreaks (10/11), with two cases re-treated with BZL due to parasite persistence.^(21,22)^ Concerning serology, data showed that IgG remains detectable for over ten years after treatment, with a low seronegative conversion rate even after re-treatment. Parasitological evaluations show an abrupt decline after six months, remain low up to two years, and stay negative thereafter. Molecular follow-up was conducted in only 3 outbreaks, two of which corresponds to the largest outbreaks reported to date, where TcI was identified.^(23,24)^ This analysis showed that the number of treated patients with detectable parasite DNA decreased progressively until year 4, with few patients regaining positivity by year 5.

**Table 5.** Serological and parasitological long-term follow-up evaluations of oral Chagas disease.

|  |  | Period evaluated |  |  |  |  |  |  |  |  |
| --- | --- | --- | --- | --- | --- | --- | --- | --- | --- | --- |
|  |  | Acute Period | < 6 months | > 6 months < 1 year | 2 years | 3 years | 4 years | 5 years | > 7 years < 8 years | > 9 years < 10 years |
| Serological follow-up | N° of outbreaks | 12 | 7 | 5 | 4 | 2 | 2 | 2 | 1 | 2 |
|  | N° of evaluated patients | 232 | 204 | 177 | 186 | 134 | 116 | 128 | 99 | 64 |
|  | IgG seropositivity (%) | 97·5 | 96·5 | 78·7 | 69·3 | 74·7 | 93·0 | 71·9 | 31·5 | 56·2 |
|  | Seronegativization (%) | 0 | 0 | 4·6 | 1·5 | 25·3 | 7·0 | n.a. | 18·6 | 28·6 |
|  | IgG seropositivity (%) - p value (vs acute period)** |  | n.s. | p<0·0001 | p<0·0001 | p<0·0001 | p=0·0455 | p<0·0001 | p<0·0001 | p<0·0001 |
| Parasitologic al follow-up | N° of outbreaks | 10 | 9 | 8 | 8 | 2 | 1 | 1 | 1 | - |
|  | N° of evaluated patients | 203 | 173 | 177 | 159 | 116 | 106 | 17 | 17 | - |
|  | Positivity (%)* | 82·7 | 6·2 | 0·12 | 0·17 | 0 | 0 | 5·9 | 0 | - |
|  | p value (vs acute period)# |  | p<0·0001 | p<0·0001 | p<0·0001 | p<0·0001 | p<0·0001 | p<0·0001 | p<0·009 | - |
| Molecular follow-up | N° of outbreaks | 3 | 1 | 1 | 2 | 1 | 1 | 1 | 1 | 2 |
|  | N° of evaluated patients | 75 | 3 | 34 | 110 | 98 | 97 | 20 | 82 | 58 |
|  | PCR Positivity (%)** | 93·0 | 33·3 | 29 | 53 | 30 | 32 | 90 | 45 | 59·5 |
|  | p value (vs acute period)# |  | p=0·02 | p<0·0001 | p<0·0001 | p<0·0001 | p<0·0001 | n.s. | p<0·0001 | p<0·0001 |

\* All parasitological methods were considered. \*\* Fisher exact test, #Chi square test.
N.a.: not available.

Food sources associated with orally transmitted ChD were reported in 63/111 outbreaks (56.7%), yet only 11 cases (10%) had reliably identified food items (IF). In the remaining events, the food source was either suspected (SCF) or not identified (NIF). Confirmed cases involved açaí (*Euterpe oleracea or E. precatoria*) and sugarcane (*Saccharum officinarum*) in 3 outbreaks each, followed by guava (*Psidium guajava*, 2/11), and individual cases involving majo fruit (*Oenocarpus bataua*), armadillo blood, and citrus juice. Suspected items included contaminated water, bacabá (*Oenocarpus bacaba*), urucuri (*Attalea phalerata*), wild animal meat, and various tropical fruits and beverages. Notably, the source remained unknown in 43·3% of outbreaks.

Açaí was most frequently linked to oral Chagas disease in Brazil (30/62), with outbreaks peaking during its harvest season, followed by an off-season from January to June.^(25,26)^ In Colombia, main sources included contaminated water, citrus juice, and armadillo meat or blood, while in Venezuela, various homemade fruit juices predominated. Notably, açaí was not reported in any Colombian or Venezuelan outbreak. Bacabá and masato were implicated in outbreaks in French Guiana and Peru, respectively. No food source was identified in the Bolivian outbreak.

The most common genera of triatomine bugs involved in food-borne ChD were *Rhodnius* (31/111; 28%) followed by *Panstrongylus* (21/111; 18·9%) and *Triatoma* (11/111; 9·9%). As seen in **Table 6**, *Rhodnius pictipes* and *Rhodnius robustus* were the most commonly identified triatomine species involved in food-borne ChD outbreaks in Brazil, whereas in Colombia and Venezuela, *Panstrongylus geniculatus* appeared as the most prevalent vector. Triatomine species involved in outbreaks from Bolivia, French Guiana, and Peru were not reported.

**Table 6:** Species of vectors suspected of being involved in food-borne ChD outbreaks.

| **Brazil**<br>[vector/ (n° outbreaks reports)] | **Colombia**<br>[vector/ (n° outbreaks reports)] | **Venezuela**<br>[vector/ (n° outbreaks reports)] |
| --- | --- | --- |
| *Rhodnius pictipes* (10)<br>*Rhodnius robustus* (9)<br>*Panstrongylus geniculatus* (3)<br>*Triatoma brasiliensis* (3)<br>*Triatoma sordida* (3)<br>*Panstrongylus lutzi* (1)<br>*Panstrongylus megistus* (1)<br>*Triatoma infestans* (2)<br>*Rhodnius brethesi* (1)<br>*Triatoma tibiamaculata* (1)<br>*Triatoma pseudomaculata* (1) | *Panstrongylus geniculatus* (8)<br>*Rhodnius pallescens* (4)<br>*Rhodnius prolixus* (3)<br>*Eratyrus mucronatus* (1)<br>*Rhodnius pictipes* (2) | *Panstrongylus geniculatus* (8)<br>*Rhodnius prolixus* (2)<br>*Triatoma maculata* (1) |

\* Data is presented as a summary of all reported species, ordered by the number of observed occurrences in each country.

*T. cruzi* is currently classified into seven DTUs (TcI–TcVI and Tcbat).^(27,28)^ TcI, TcIII, and TcIV are considered mainly sylvatic, while TcII, TcV, and TcVI are primarily domestic.^(29)^ Molecular data from 38 outbreaks (33·9%) revealed DTU involvement, with sylvatic lineages present in 97·4% (37/38). In 6 of 7 outbreaks with confirmed food sources, TcI (2 outbreaks) or TcIV (1 outbreak) were identified by qPCR in both food and human samples. In the other, the identification was only performed in human samples and also corresponded to TcI and TcIV. In the 31 remaining outbreaks, TcI was the most reported, followed by TcIV. TcII was detected in 2 outbreaks. Moreover, mixed DTUs, such as TcIII/TcIV (3 outbreaks) or TcI/TcII/TcVI (1 outbreak) were also associated with oral infection. In 12 outbreaks, TcI was also identified in triatomines collected in the area. The geographic distribution of identified DTUs are shown in **Figure 6**.

**Figure 6:**
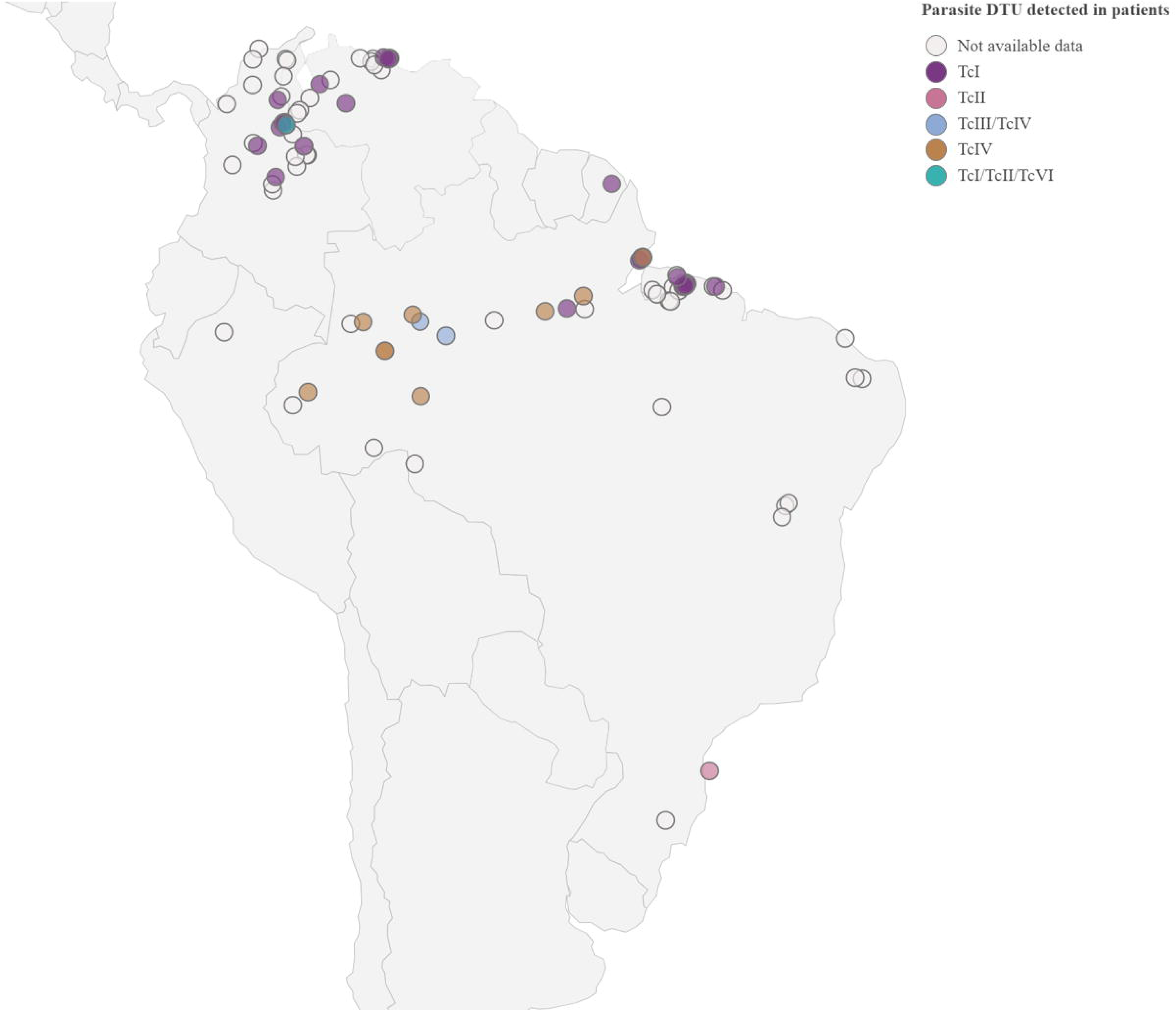
Geographic distribution of DTUs associated with food-borne Chagas disease outbreaks.

Lastly, ***c***lassification of food-borne outbreaks according to quality and quantity of extractable data showed that only 10 outbreaks (9%) were reported with highly detailed data. Among them, only one study had an unequivocal detection of the parasite in the contaminated food. Additionally, 31 outbreaks (28%) were described with intermediate detailed data, while the 70 rest (63·1%) had limited (47·7%) or scarce (15·3%) description.

## DISCUSSION

Food-borne transmission accounts for nearly 15% of human emergent infectious disease events, with a significant proportion caused by protozoan and helminth parasites.^(30)^ Acute food-borne *T. cruzi* infections are increasingly reported across Latin America, yet often go undiagnosed and untreated due to limited recognition and resources.^(31)^ Despite growing concern, key aspects of this transmission route remain poorly defined, and few systematic studies have addressed its specific features.^(4,32)^ Our study addresses this gap through a comprehensive analysis of scientific and grey literature from the past six decades, offering an expanded view of outbreak characteristics and impact.

Food-borne Chagas disease (ChD) outbreaks are typically point-source events affecting small to medium-sized groups, with a median incubation of 16 days.^(33)^ This delay complicates the identification of contaminated sources due to recall bias and delayed epidemiological surveys. Effective tracing requires preserving suspected food items for laboratory analysis—yet, as shown in this review, proper storage enabling source identification occurred only once. Detection is further hindered by inadequate sampling, environmental factors, and logistical barriers.^(34,35)^ These challenges limit outbreak control and underscore the need for improved surveillance, food safety protocols, and One Health-based protocols to better approach this issue. In particular, foods like açaí and sugar cane in Brazil, and diverse fruits in Colombia and Venezuela, require more stringent safety standards to reduce the risk of contamination.

Our study confirms a seasonal pattern in food-borne Chagas disease outbreaks, with higher incidence during warm and rainy months, coinciding with harvest seasons and increased triatomine activity in peridomestic areas.^(36)^ These findings highlight the importance of incorporating seasonal dynamics into *T. cruzi* surveillance and adopting One Health-based strategies. Factors such as deforestation, crop harvesting, and urban expansion—often accompanied by waste, water, or food accumulation—may facilitate interactions between sylvatic vectors and domestic environments, enhancing the risk of food-borne transmission.^(34,37)^ This is supported by the predominance of sylvatic *T. cruzi* DTUs in the outbreaks analyzed, accounting for 98% of reported cases. In addition, parasitological assessments confirm that early treatment of oral Chagas disease rapidly reduces parasitemia and mitigates acute symptoms. However, genetic variability among *T. cruzi* strains—particularly TcI, which shows some drug resistance in vitro and in clinical cases, highlights the need for alternative treatment regimens. ^(16,38–40)^

Additionally, the oral route influences disease severity, including acute symptoms, cardiomyopathy progression, and lethality. In this review, the lethality rate was 6.5% (range: 0–100%), higher than the 1% (95% CI: 0.0–4.0%) reported in a meta-analysis of 2,470 cases^(32)^, and markedly above the 0.2–0.5% for vector-borne infections^(33)^, though slightly below earlier estimates of 8–35%.^(2)^ With 77 deaths linked to oral outbreaks and limited cardiological data, severe cardiac complications such as heart failure or cardiogenic shock may be underestimated.

Our study highlights the critical lack of long-term follow-up data on oral Chagas disease, limiting insight into its potential impact on chronic cardiac and gastrointestinal complications, even in treated patients. While parasite genetic diversity may influence disease progression, symptoms, and treatment response, the scarcity of long-term molecular and clinical studies hampers our understanding of its role in treatment failure and outcomes. These gaps underscore the urgent need for targeted research.^(41,42)^

Key strengths of this study include a comprehensive literature search across major databases (PubMed, Embase, Web of Science, and Scielo) and the inclusion of gray literature such as national health reports, PAHO documents, and academic theses, enhancing data coverage and minimizing the risk of missed reports. Sources in English, Portuguese, and Spanish were reviewed. However, limitations were evident, particularly due to resource constraints affecting data collection and the potential for publication bias from unreported outbreaks or missing variables. Challenges during data extraction included incomplete or missing information (e.g., sex, age, latency, outbreak dates, number of patients with a determinate signs or symptoms), variable data quality, limited supporting evidence for some findings, and significant heterogeneity— especially between oral and vector-borne cases. Patient-specific factors like ethnicity and comorbidities, as well as inconsistencies across reports of the same outbreak, can significantly contribute to the heterogeneity of study results.

### Final remarks

Over recent decades, our understanding of ChD has expanded, with increasing recognition of its global distribution driven by migration and the growing awareness of food-borne transmission in Latin America. Effectively addressing its prevention, management, and monitoring requires a multidisciplinary approach that integrates clinical care, epidemiological surveillance, and basic research. Despite the clear impact of food-borne transmission, many regions, including parts of the Southern Cone (e.g., Argentina and Chile), Mexico, and Central America, report little or no data on oral ChD, suggesting a potential underdiagnosis.

While most outbreaks have been described in terms of epidemiology and clinical presentation, the source of contamination and the specific vectors involved are often not identified. This limits our understanding of transmission dynamics and highlights the need for a more integrated approach to studying food-borne transmission of ChD, considering ecological factors, diagnostic and treatment challenges, and the importance of enhanced epidemiological surveillance. It also stresses the need for greater public awareness of the risk of infection, especially when people are simultaneously exposed to the same transmission. Our review underscores the importance of a more integrated, One Health-oriented approach to food- borne ChD, accounting for ecological, diagnostic, and public health challenges. Public awareness must also be strengthened, particularly in scenarios of group exposure to contaminated food or water. By systematically analyzing epidemiological and clinical data from 111 outbreaks reported over six decades in South America, we identified significant knowledge gaps and the urgent need for standardized evidence to inform better control strategies. Filling these gaps is essential to improve clinical management and surveillance, especially in endemic regions. Furthermore, advancing research on host immune responses and the biology of oral transmission will be critical for developing targeted prevention measures. Our findings provide a foundation to guide future investigations and reporting practices. Ultimately, this work calls for coordinated action by national and international health agencies to define a global protocol that ensures the systematic and reliable documentation of oral ChD outbreaks.

## Supporting information

Supplemental File - PRISMA

## Data Availability

The data supporting the findings of this review are available at RDA-UNR: https://doi.org/10.57715/UNR/ZFRC4G

## Author Contributions

**UYM, Dinatale B, Bottasso O, Pérez AR, Savino W.** All authors had full access to all the data in the study and had a final responsibility for the decision to submit for publication.

## Data sharing

The data supporting the findings of this review are available at: https://doi.org/10.57715/UNR/ZFRC4G

