## Supplemental File - PRISMA for "Outbreaks of orally transmitted Chagas disease in Latin America: a comprehensive systematic review"

### Useche YM et al, 2025. PRISMA checklist

| Section and Topic | Item # | Items and elements recommended for reporting |
|---|---|---|
| **TITLE** | **1** | ***Identify the report as a systematic review***<br><br>**Outbreaks of orally transmitted Chagas disease in Latin America: a comprehensive systematic review** |
| **ABSTRACT** | | |
| **ABSTRACT** | **2** | The abstract has been prepared in accordance with the PRISMA 2020 for Abstracts checklist, addressing the corresponding items |
| **INTRODUCTION** | | |
| **RATIONALE** | **3** | ***Rationale for the review in the context of existing knowledge***<br><br>The rising burden of orally transmitted Chagas disease in South America underscores the need for a deeper understanding of this food-borne transmission route. Despite its relevance, key aspects such as transmission dynamics, epidemiology, and clinical outcomes remain insufficiently explored, limiting prevention and treatment strategies. |
| **OBJECTIVES** | **4** | ***Objectives or questions the review addresses***<br><br>This systematic review aims to characterize oral Chagas disease outbreaks to provide comprehensive evidence on the symptomatology, diagnosis, and treatment of patients with oral infections through a systematic review of observational studies conducted between 1965 and 2023 across Latin America. It seeks to provide comprehensive evidence on the symptomatology, diagnosis, and treatment of orally infected patients, as well as clinical and serological follow-up outcomes. Additionally, it compiles data on contaminated sources, vector species, parasite lineages, and seasonality. |
| **METHODS** | | |
| **ELIGIBILITY CRITERIA** | **5** | ***Inclusion and exclusion criteria for the systematic review***<br><br>Eligible reports were those describing oral outbreaks of Chagas disease. Longitudinal, ecological, in vitro, animal, or non–Chagas-related parasitic studies were excluded. Full texts were reviewed when abstracts were inconclusive. Studies lacking diagnostic confirmation, evidence of oral infection, or extractable data were excluded. Disagreements were resolved following the Cochrane Handbook. Data discrepancies were discussed and checked against the original sources.<br><br>Studies or reports corresponding to oral Chagas outbreaks occurred between 1965 and 2023 in Latin America countries were included. No language limits were applied. Gray literature sources were searched to identify possible relevant data about outbreaks descriptions not previously included in peer-reviewed scientific publications. These sources included PhD thesis, dissertation databases, conference abstracts, and online local newspapers reports. We also included reported cases of acute ChD from Colombian and Brazilian online government health surveillance databases. |

| | | |
|---|---|---|
| **INFORMATION SOURCES and SEARCH STRATEGY** | **6,7** | ***List of sources used to identify studies (including databases, registers, websites or organizations), along with full search strategies.***<br><br>This systematic review followed the PRISMA 2020 statement, and is based on a protocol (PROSPERO 2024 CRD42024542461) registered prospectively on 7 June 2024. At first, there are no restrictions on the types of study included. Period: 01/01/1950 until 12/31/2023.<br>An electronic search for the available literature (database inception to 28 December 2023) was performed using Pubmed, Web of Science, Embase, LILACS, ResearchGate and Scielo databases including the followed DeCS/MeSH terms: "Chagas Disease", "*Trypanosoma cruzi Infection*", "foodborne diseases", "outbreak", and other terms related to oral transmission by contaminated food like: "contaminated food", "oral infection", "oral transmission" and several synonyms were searched in titles and abstracts. Truncation was used to ensure variant terms were retrieved. The references were exported to EndNote X7.5 and transferred to the Rayyan (https://www.rayyan.ai/) online application for the selection process. |
| **SELECTION PROCESS** | **8** | ***Study selection process, including number of reviewers, their level of independence, and any automation tools used.***<br><br>Three authors (YMU, ARP, BD) screened titles and abstracts based on inclusion/exclusion criteria, including DeCS/MeSH terms. Disagreements were resolved by a fourth author (OB), following the Cochrane Handbook. Data discrepancies were discussed and checked against the original sources. |
| **DATA COLLECTION PROCESS** | **9** | ***Methods used to collect data from reports, including how many reviewers collected data from each report, whether they worked independently, and if applicable, details of automation tools used in the process.***<br><br>Data from the included studies were independently and thoroughly collected and extracted by 3 persons using an Excel form (ARP, YMU, BD). Data was extracted and entered into an Excel sheet and disagreement will be resolved by discussion with a fourth reviewer (OB). No automation tools were used during the process. |
| **DATA ITEMS** | **10** | ***List of data and/or outcomes collected.***<br><br>Characteristics of included studies be summarized as tables. The following information will be extracted: type of study, first author, temporal and geographical data (month/year and country of outbreaks), involved people numbers (outbreak size), age and sex of infected patients, pregnant women who have abortions during acute infection, diagnosis methods, disease signs and symptoms, case-fatality rates, treatments, clinical and serological follow-up studies, food-borne source and involved T. cruzi DTUs. |
| **STUDY RISK OF BIAS ASSESSMENT** | **11** | Review authors will independently assess the quality of each included record. In addition, a classification of food-borne outbreaks according to quality and quantity of extractable data was performed. Outbreaks were categorized by the certainty level of available data in high, intermediate, limited or scarce. |
| **EFFECT MEASURES** | **12** | ***Measures used in the synthesis or presentation of results.***<br><br>Quantitative variables were summarized as means with standard deviations for normally distributed data, or as medians with ranges or quartiles for non-normally distributed data. Qualitative (categorical) variables were described using absolute and relative frequencies (percentages). |
| **SYNTHESIS METHODS** | **13** | ***Synthesis Methods used in the Systematic Review***<br><br>Normality was assessed using the Shapiro-Wilk test. Categorical variables were compared using the chi-square test or Fisher's exact test when appropriate, while continuous variables were analyzed using Student's t-test or the Mann-Whitney U test, depending on data distribution. Descriptive statistics were also presented graphically, as appropriate.The geographical distribution map was created using Scimago Graphica (https://www.graphica.app/), incorporating geolocation data |

| | | |
|---|---|---|
| | | obtained from bibliographic sources or Google Maps. |
| **REPORTING BIAS ASSESSMENT** | **14** | Not applicable |
| **CERTAINTY ASSESSMENT** | **15** | Not applicable |
| **RESULTS** | | |
| **STUDY SELECTION** | **16** | ***Flow of studies*** A flow diagram was created to describe the number of records identified, including observational studies and grey literature. The number of reports remaining after automatic duplicate removal and those excluded due to ineligibility were documented. Publications excluded due to the absence of extractable data or information on positive diagnostic tests were also reported. The total number of reports and cases included in this systematic review is presented in the flow diagram. |
| **STUDY CHARACTERISTICS** | **17** | ***Study characteristics*** The database search including observational studies as outbreak reports, outbreak series or reviews with details of outbreak series, as well as studies of molecular typing of the strains involved in the outbreaks and grey literature articles including theses, conference abstracts, governmental health agency reports, and online journals, while press reports were included to analyze seasonality. |
| **RISK OF BIAS IN STUDIES** | **18** | Not applicable |
| **RESULTS OF INDIVIDUAL STUDIES** | **19** | Not applicable |
| **RESULTS OF SYNTHESES** | **20** | We compiled data from 111 outbreaks involving 1187 cases (55% males), 77 deaths, in 6 countries. Contaminated food sources were identified in 63 outbreaks (56·7%), with reliable confirmation in 8 (7·2%), *açaí* fruit being the most common source (28%). Outbreaks occurred mainly in sylvatic and tropical regions, coinciding with warm seasons and crop harvest periods. Selvatic parasite lineages predominated (98%), with *Rhodnius spp*. triatomines being implicated in 28%. The median incubation period was 22 days. Case fatality ratio was 6·5%. Acute infection presented with fever (80%), facial edema (20%), altered ECG (39·2%) and Echo (28·9%). Despite antiparasitic treatment, cardiac alterations persisted after 1 and 4 years. A ten-year follow-up showed no chronic Chagasic cardiomyopathy, though risk markers and persistent *T. cruzi*-specific IgG were reported. Data reinforces gaps mainly in long-term clinical, serological and molecular follow-up. |

| | | |
|---|---|---|
| **REPORTING BIASES** | **21** | Not applicable |
| **CERTAINTY OF EVIDENCE** | **22** | Not applicable |
| **DISCUSSION** | | |
| **DISCUSSION** | **23** | • Discussion provide a general interpretation of the results and discuss limitations of the systematic review. Moreover, include implications of the results for clinical practice and government health policy.Make explicit recommendations for future research on food-borne Chagas disease. |
| **OTHER INFORMATION** | | |
| **REGISTRATION AND PROTOCOL** | **24** | ***Registration:*** <br> This systematic review followed the PRISMA 2020 statement, and is based on a protocol (PROSPERO 2024 CRD42024542461) registered prospectively on 7 June 2024. At first, there are no restrictions on the types of study included. Period: 01/01/1950 until 12/31/2023. |
| **SUPPORT** | **25** | ***Sources of financial or non-financial support for the review*** <br> Review has no specific/external funding but is supported by guarantor/review team (non-commercial) institutions |
| **COMPETING INTERESTS** | **26** | ***Declaration*** <br> Authors had no competing interests. |
| **AVAILABILITY OF DATA, CODE, AND OTHER MATERIALS** | **27** | ***Repository*** <br> Data extracted and used for all analyses will be available at the Universidad Nacional de Rosario public repository. https://rephip.unr.edu.ar/home <br> Other public repository could be also contemplated. |

*This PRISMA Checklist is based on:* Page MJ, McKenzie JE, Bossuyt PM, Boutron I, Hoffmann TC, Mulrow CD, et al. The PRISMA 2020 statement: an updated guideline for reporting systematic reviews. BMJ

2021;372:n71. doi: 10.1136/bmj.n71.
